# Clinical Research Collaboration for Stroke in Korea Imaging Repository: A Prospective Multicenter Neuroimaging Repository

**DOI:** 10.64898/2026.03.17.26348664

**Authors:** Beom Joon Kim, Wi-Sun Ryu, Myung Jae Lee, Kyusik Kang, Jae Guk Kim, Soo Joo Lee, Jae-Kwan Cha, Tai Hwan Park, Jeong-Yoon Lee, Kyungbok Lee, Doo Hyuk Kwon, Jun Lee, Hong-Kyun Park, Yong-Jin Cho, Keun-Sik Hong, Minwoo Lee, Mi Sun Oh, Kyung-Ho Yu, Dong-Seok Gwak, Dong-Eog Kim, Hyunsoo Kim, Joon-Tae Kim, Joong-Goo Kim, Jay Chol Choi, Wook-Joo Kim, Young Cheol Weon, Jee Hyun Kwon, Kyu Sun Yum, Dong-Ick Shin, Jeong-Ho Hong, Sung-Il Sohn, Sang-Hwa Lee, Chulho Kim, Hae-Bong Jeong, Kwang-Yeol Park, Chi Kyung Kim, Jihoon Kang, Jun Yup Kim, Do-Yeon Kim, Jongwuk Kim, Nakhoon Kim, Bijoy K. Menon, Longting Lin, Mark Parsons, Hee-Joon Bae

## Abstract

**Background:** Prospective stroke registries have advanced our understanding of cerebrovascular disease, yet most reduce neuroimaging to categorical variables, forfeiting the multidimensional information inherent in clinical imaging. We describe the CRCS-K Imaging Repository, a prospective multicenter platform that systematically collects all stroke neuroimaging and integrates artificial intelligence (AI)-based automated quantification with clinical and outcome data through a dedicated research platform, AISCAN.

**Methods:** Building upon the Clinical Research Collaboration for Stroke in Korea (CRCS-K), a nationwide prospective registry, all neuroimaging (computed tomography [CT], magnetic resonance [MR], and angiography) performed during index hospitalization of consecutive acute ischemic stroke patients was collected from 18 comprehensive stroke centers. Imaging underwent centralized quality verification, sequence classification, and AI-based quantification. As a proof-of-concept application, we examined the association between pre-treatment imaging modality, treatment workflow efficiency, and functional outcomes in patients receiving intravenous thrombolysis (IVT) or endovascular treatment (EVT).

**Results:** From June 2022 through May 2025, 225,159 imaging sequences were collected from 20,792 patients. AI-based quantification modules converted these into standardized numeric features encompassing ischemic lesion volumes, perfusion parameters, white matter hyperintensity burden, and cerebral microbleed counts. Substantial inter-hospital variation in imaging modality selection was observed, with MR-first workflows ranging from 1.0% to 56.7% across centers. In the proof-of-concept analysis, each additional imaging sequence was associated with prolonged door-to-treatment times for both IVT and EVT. Propensity score overlap-weighted analyses suggested numerically more favorable functional outcomes with CT-based imaging among EVT-treated patients, whereas differences among IVT-treated patients were smaller and less consistent.

**Conclusions:** The CRCS-K Imaging Repository demonstrates the feasibility of large-scale, prospective neuroimaging collection integrated with AI-based quantification and clinical data. The infrastructure enables clinically consequential questions that conventional registries cannot address.

## Introduction

Neuroimaging occupies a central role in acute ischemic stroke management, informing both recanalization decisions and long-term therapeutic strategies.^1^ Yet the translation of imaging information into large-scale research remains constrained by a fundamental tension: whereas randomized controlled trials employ standardized protocols in carefully selected populations,^2^ observational stroke registries, which capture the breadth of real-world practice, typically reduce neuroimaging to a circumscribed set of categorical variables such as the site of arterial occlusion or semi-quantitative grading of small vessel disease.^3–5^ This inevitable simplification forfeits the multidimensional information inherent in multimodal and serially acquired imaging studies. Recent advances in computer sciences, artificial intelligence (AI) and automated image analysis offer the prospect of storing a large mass of images and converting those into standardized quantitative metrics at scale,^6–8^ yet robust development and validation of such tools demand large volumes of imaging data tightly coupled with clinical information, maintained longitudinally across diverse centers and temporal epochs.^9,10^ Several imaging-enriched stroke registries have recently emerged^11,12^ yet none systematically integrate the prospective collection of all neuroimaging modalities with AI-based automated quantification.

The Clinical Research Collaboration for Stroke in Korea (CRCS-K) has maintained a nationwide prospective registry of acute stroke patients since 2008, with systematic collection of clinical data and ascertainment of functional and event outcomes.^13,14^ Building upon this infrastructure, the authors established the CRCS-K Imaging Repository, a prospective multicenter platform that systematically collects all neuroimaging performed during the index hospitalization for acute ischemic stroke and integrates AI-based automated quantification with clinical and outcome data. In the present paper, we describe the development and structure of this repository, summarize the characteristics of the accumulated imaging and clinical data, introduce an integrated research platform AISCAN, and, as a proof-of-concept application illustrating the research utility of this infrastructure, investigate the association between pre-treatment neuroimaging workflow and the efficiency and effectiveness of intravenous thrombolysis (IVT) and endovascular treatment (EVT) in real-world clinical practice.

## Methods

The data that support the findings of this study are available from the corresponding author upon reasonable request.

### Target Population

The CRCS-K Imaging Repository was established in 2022, building upon CRCS-K, a prospective multicenter nationwide clinical stroke registry. Participating hospitals voluntarily joined the Imaging Repository according to their institutional capacity and resources. The CRCS-K registry enrolls patients presenting with acute ischemic stroke or transient ischemic attack within 7 days of symptom onset; enrollment requires written informed consent from the patient or a legally authorized representative for the registration of clinical data, imaging, and biosignals. The CRCS-K Imaging Repository applies identical enrollment criteria. Initial patient registration commenced in the latter half of 2022 and was subsequently expanded across the full CRCS-K registry network, ultimately encompassing 18 hospitals throughout the Republic of Korea. The study was approved by the institutional ethics committees of the central coordinating center, Seoul National University Bundang Hospital (IRB Approval No. B-1706-403-303 and B-2307-841-303), and all participating institutions.^13,14^

### Neuroimaging Data Collection Process

Clinical management of patients with acute ischemic stroke at each participating hospital was conducted in accordance with contemporary clinical practice guidelines and institution-specific protocols. Accordingly, the selection of neuroimaging modality for suspected acute stroke patients in the emergency department and the determination of follow-up neuroimaging varied considerably across institutions. The Imaging Repository collected all stroke neuroimaging studies performed from the time of hospital arrival through discharge or transfer to a rehabilitation facility during the index hospitalization, encompassing computed tomography (CT), magnetic resonance (MR) imaging, and digital subtraction angiography (DSA). Ultrasonographic studies were excluded owing to substantial heterogeneity in institutional protocols. All neuroimaging data were collected in native Digital Imaging and Communications in Medicine (DICOM) format with header information preserved. During this process, all personally identifiable information was removed, and each study was assigned a pseudonymized identifier linked to the CRCS-K registry.

As a prospective clinical stroke registry, CRCS-K requires initial registration of each patient within 24 business hours of hospital admission, with subsequent entry of detailed clinical information. Three-month modified Rankin Scale (mRS) scores and event outcome data are collected thereafter, and the verified dataset is released to investigators. Imaging data collection for the CRCS-K Imaging Repository commences after the release of each verified clinical dataset. Each participating hospital then compiles and transmits anonymized neuroimaging data to the central imaging laboratory. The central laboratory subsequently conducts quality verification according to a standardized protocol, assessing patient completeness, imaging completeness, and data integrity. Discrepancies prompt issuance of formal queries for re-collection or correction. Upon verification, systematic data extraction from the imaging repository is initiated.

### Data Extraction, Image Cataloguing, and AI-Based Quantification

All imaging data undergo quality verification at the central imaging laboratory, where removal of personally identifiable information is confirmed and pseudonymized identifiers are assigned. Essential metadata are extracted from the neuroimaging files during this process. Pseudonymized imaging data and associated metadata are subsequently transmitted to the central imaging server hosted at the CRCS-K Imaging Repository through a secure, encrypted channel. Discrepancies and corrupted files are regularly queried back to the local site for resolution.

The central imaging server catalogues each imaging study on the basis of the CRCS-K unique identifier, participating center, imaging date and time, and modality. Predefined sets of AI-based quantification models, validated in multicenter studies,^15–22^ are then applied to eligible images, and the resulting numeric outputs are appended to the integrated dataset. Detailed listings of the deployed AI modules at the time of the current publication are provided in Supplemental Table S1.

### AISCAN: Versatile and Scalable Research Platform

Imaging data and AI-derived quantified neuroimaging features collected through the CRCS-K Imaging Repository, together with clinical information collected through the CRCS-K registry, are disseminated to qualified researchers through AISCAN (https://aiscan.medihub.ai), a versatile and scalable web-based platform that integrates clinical information and imaging data to facilitate clinical research. AISCAN presents core variables derived from the CRCS-K registry, including demographics, vascular risk factors, stroke subtype, acute treatment, temporal indicators, in-hospital management, and event and functional outcomes, alongside each patient’s imaging data containing metadata and AI-derived quantitative features. Clinical and imaging data tables are linked through pseudonymized CRCS-K unique identifiers, and the platform incorporates comprehensive query and filtering capabilities across all data fields, enabling researchers to rapidly identify and review study subjects pertinent to their individual research questions. AISCAN is architected to be both scalable and versatile, enabling the incorporation of newly developed AI modules, additional imaging modalities, and external clinical datasets with minimal modifications to the core infrastructure. Detailed technical specification and data security architecture of the AISCAN platform were provided as Supplemental Data 1 and 2.

Access to AISCAN is granted through a formal application process, with proposals reviewed and approved by the CRCS-K Imaging Repository steering committee. External investigators submit a brief project proposal outlining the research question, requested variables, selection criteria, and analysis plan. Upon approval, the AISCAN data manager constructs a project-specific dataset, and the investigator is granted access restricted to that particular dataset. All operations are logged, and data are accessible only within the secure environment; export of imaging and clinical data is restricted to aggregate or anonymized outputs in accordance with applicable regulations of the Republic of Korea and relevant jurisdictions.

### Data Ownership, Access, and Publication

Each participating center retains primary ownership of its own clinical and imaging data. The central coordinating center of the CRCS-K Imaging Repository acts as custodian of the collected data and the merged registry-imaging dataset. Access to the integrated data is granted on a project-by-project basis following review and approval of a written proposal by the CRCS-K Imaging Repository steering committee. For each approved project, participating centers may elect whether to contribute their data.

### Statistical Analysis of Imaging Strategy and Treatment Workflow Metrics

To demonstrate the research utility of the integrated clinical and imaging dataset, we examined how the modality and sequence composition of acute stroke neuroimaging used for initial patient triage influenced the efficiency and effectiveness of acute recanalization treatment. For this analysis, all imaging acquired within three hours of hospital arrival was provisionally designated as initial triaging imaging. In patients who received IVT or EVT, all imaging performed prior to treatment initiation was classified as acute stroke neuroimaging. When both CT-based and MR-based sequences were obtained, the patient was classified as having undergone MR-based decision-making. Patients were categorized into the following imaging protocol groups: pre-arrival imaging only, CT-based protocols (non-contrast CT [NCCT] only, CT angiography [CTA] with or without NCCT, or CT perfusion [CTP] plus CTA with or without NCCT), and MR-based protocols (MR with or without perfusion imaging). Across all participating hospitals, the stroke MR protocol included at minimum diffusion-weighted imaging (DWI) and intracranial MR angiography.

Continuous variables were expressed as means ± standard deviations or medians [interquartile ranges], and categorical variables as counts (percentages). Between-group comparisons employed appropriate parametric or non-parametric tests. To assess the association between imaging strategy and treatment time metrics, we fitted multivariable hierarchical mixed-effects models with random intercepts for centers, adjusting for relevant covariates. For the workflow efficiency analysis, linear mixed-effects models were applied to log-transformed door-to-needle (for IVT) or door-to-puncture (for EVT) times to accommodate right-skewed distributions. These models were adjusted for age, sex, onset-to-arrival time, baseline National Institutes of Health Stroke Scale (NIHSS) score, stroke etiology, vascular risk factors, last-known-well to arrival time, and arrival year-month, with hospital included as a random intercept; EVT models were additionally adjusted for bridging thrombolysis.^23^ For the mRS shift analysis, we used cumulative link mixed models with a logit link and a random intercept for site to account for center-level clustering, adjusting for age, sex, baseline NIHSS, pre-stroke mRS, and comorbidities.

To evaluate the association between triaging modality and treatment outcome, we implemented propensity score analyses employing both inverse probability of treatment weighting (IPTW) and overlap weighting (OW), the latter selected to focus inference on patients with substantial treatment overlap, particularly in the presence of asymmetric propensity score distributions. Balance was assessed via standardized mean differences (SMD), with a threshold of <0.10 deemed acceptable; any covariates failing to achieve this threshold were additionally included in the outcome model. Functional outcomes were assessed as the mRS score at 3 months, dichotomized at 0 to 1 and 0 to 2, as well as ordinal shift analysis with cluster-robust standard errors. Treatment effects were estimated using doubly robust weighted logistic regression, and marginal risk differences and risk ratios were derived with 95% confidence intervals calculated via cluster-bootstrap resampling over 1,000 iterations.^24^

A two-tailed P-value of <0.05 was considered statistically significant. All statistical analyses were performed using R version 4.5.2 (R Foundation for Statistical Computing, Vienna, Austria) with the *lme4* package (version 1.1-38).

## Results

### Patient Enrollment and Neuroimaging Data Collection

Over the three-year period from June 2022 through May 2025, a total of 20,792 patients from 18 comprehensive stroke centers throughout the Republic of Korea were enrolled in the CRCS-K Imaging Repository, with both clinical and neuroimaging data collected (Figure 1). Among enrolled patients, 12,384 (59.6%) were male, the mean age was 69.6 ± 13.5 years, and the median baseline NIHSS score was 3 [interquartile range (IQR), 1 to 7]. The median time from last known well to hospital arrival was 11.3 hours [IQR, 2.8 to 33.4]. Recanalization treatment was administered in 3,612 patients (17.4%), comprising 2,258 (10.9%) who received IVT and 2,173 (10.5%) who received EVT (Table 1).

**Figure 1.**
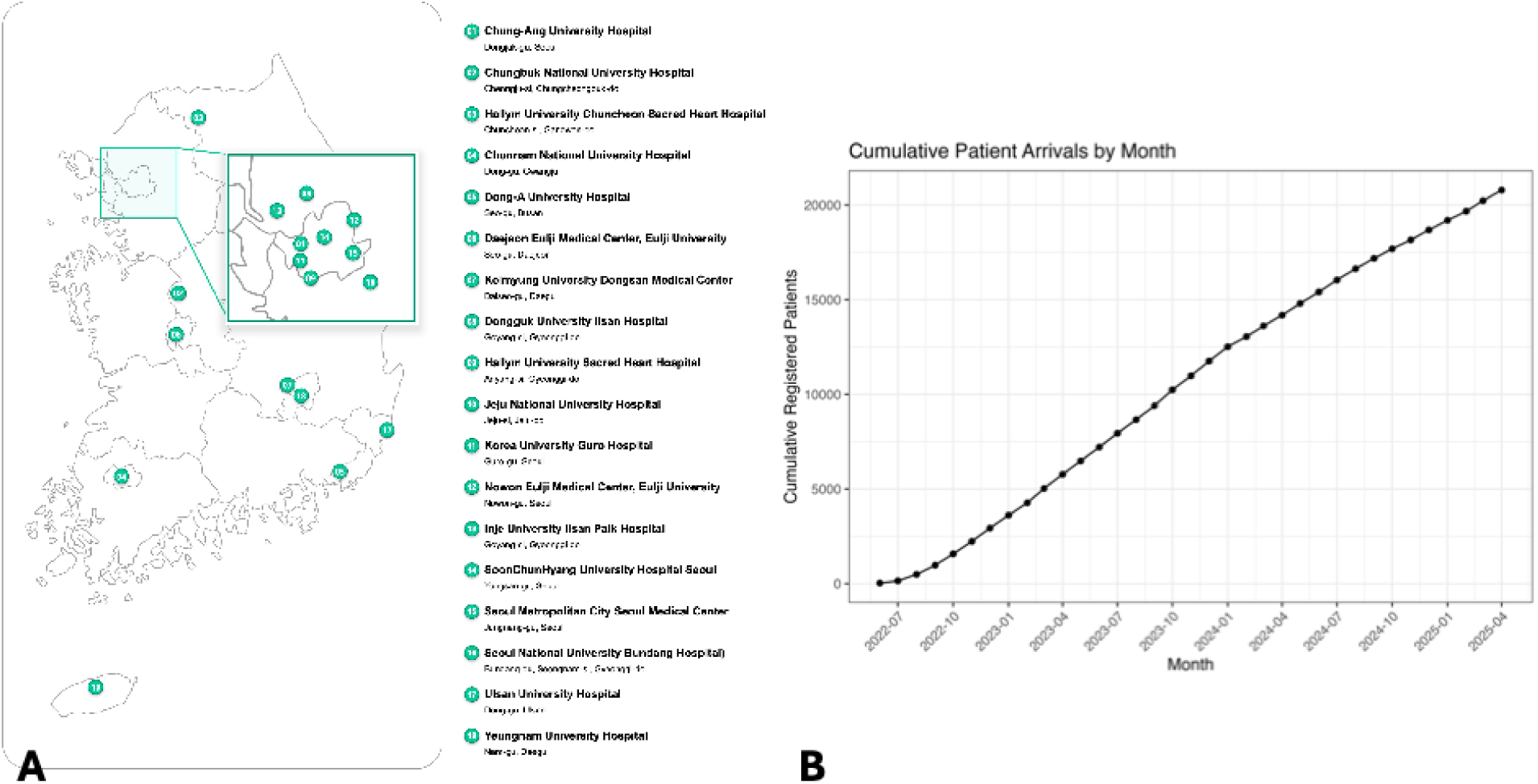
Participating Centers and Temporal Progression of the CRCS-K Imaging Repository. (A) Geographic distribution of the 18 participating comprehensive stroke centers across the Republic of Korea. (B) Cumulative patient enrollment and imaging registration by calendar month from June 2022 through May 2025.

**Table 1.**
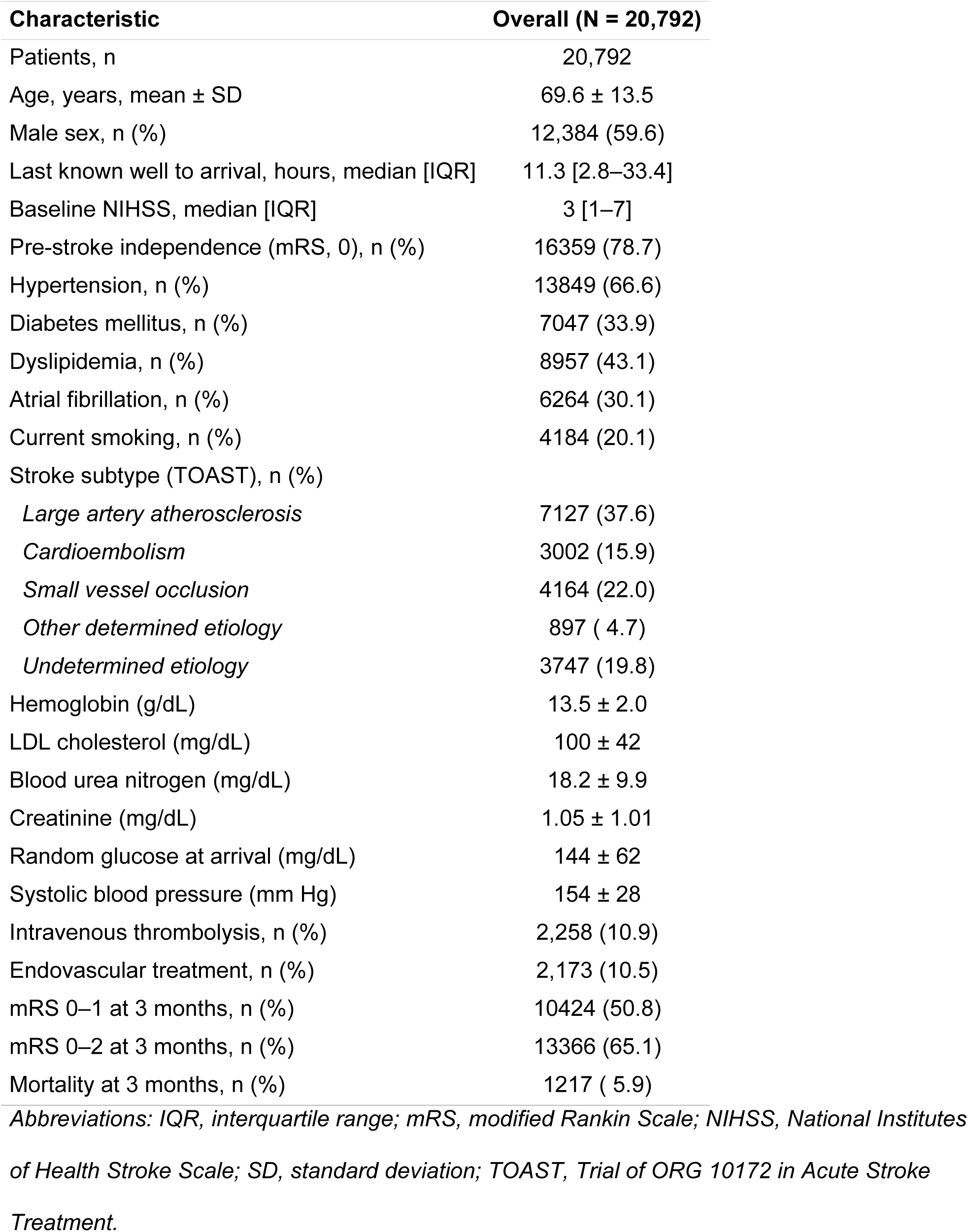
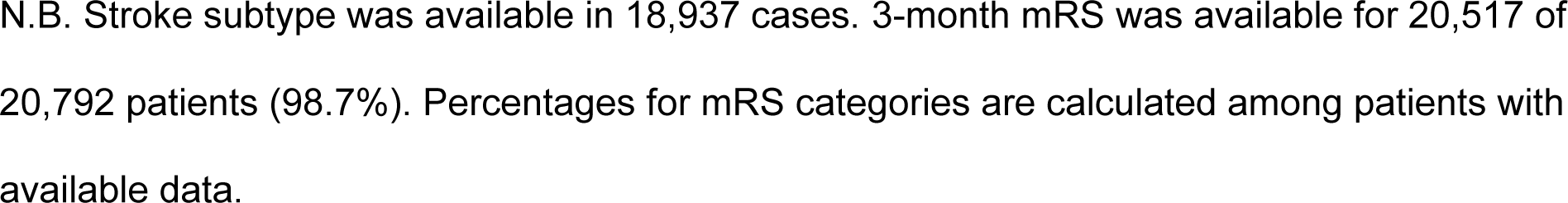
Baseline Characteristics of Patients Enrolled in the CRCS-K Imaging Repository.

| Characteristic | Overall (N = 20,792) |
| --- | --- |
| Patients, n | 20,792 |
| Age, years, mean ± SD | 69.6 ± 13.5 |
| Male sex, n (%) | 12,384 (59.6) |
| Last known well to arrival, hours, median [IQR] | 11.3 [2.8–33.4] |
| Baseline NIHSS, median [IQR] | 3 [1–7] |
| Pre-stroke independence (mRS, 0), n (%) | 16359 (78.7) |
| Hypertension, n (%) | 13849 (66.6) |
| Diabetes mellitus, n (%) | 7047 (33.9) |
| Dyslipidemia, n (%) | 8957 (43.1) |
| Atrial fibrillation, n (%) | 6264 (30.1) |
| Current smoking, n (%) | 4184 (20.1) |
| Stroke subtype (TOAST), n (%) |  |
| *Large artery atherosclerosis* | 7127 (37.6) |
| *Cardioembolism* | 3002 (15.9) |
| *Small vessel occlusion* | 4164 (22.0) |
| *Other determined etiology* | 897 ( 4.7) |
| *Undetermined etiology* | 3747 (19.8) |
| Hemoglobin (g/dL) | 13.5 ± 2.0 |
| LDL cholesterol (mg/dL) | 100 ± 42 |
| Blood urea nitrogen (mg/dL) | 18.2 ± 9.9 |
| Creatinine (mg/dL) | 1.05 ± 1.01 |
| Random glucose at arrival (mg/dL) | 144 ± 62 |
| Systolic blood pressure (mm Hg) | 154 ± 28 |
| Intravenous thrombolysis, n (%) | 2,258 (10.9) |
| Endovascular treatment, n (%) | 2,173 (10.5) |
| mRS 0–1 at 3 months, n (%) | 10424 (50.8) |
| mRS 0–2 at 3 months, n (%) | 13366 (65.1) |
| Mortality at 3 months, n (%) | 1217 ( 5.9) |

*Abbreviations: IQR, interquartile range; mRS, modified Rankin Scale; NIHSS, National Institutes* *of Health Stroke Scale; SD, standard deviation; TOAST, Trial of ORG 10172 in Acute Stroke* *Treatment.*
N.B. Stroke subtype was available in 18,937 cases. 3-month mRS was available for 20,517 of 20,792 patients (98.7%). Percentages for mRS categories are calculated among patients with available data.

A total of 225,159 imaging sequences were collected in the repository, including 37,453 DWI, 35,242 NCCT, 24,829 time of flight MR angiography, 20,508 susceptibility-weighted imaging, 16,729 gradient-echo imaging, 15,503 CTA, 4193 CTP, and 5809 MR perfusion-weighted imaging (PWI) sequences, among others (Supplemental Table S2). The mean number of DWI sequences per imaged patient was 1.85 ± 0.83, and the mean number of NCCT acquisitions was 2.04 ± 1.96. Quantified imaging features derived from these sequences through the AI-based pipeline are detailed in the Supplemental Data 3. A total of 2,906 patients (14.0%) had undergone stroke imaging at an outside facility prior to arrival at a CRCS-K participating hospital; among these, DWI had been performed in 81.6% and NCCT in 55.8% (Supplemental Table S3).

### Inter-Hospital Heterogeneity in Acute Stroke Neuroimaging Modality

Substantial inter-hospital variation was observed in the imaging modality employed for acute stroke triage among the 18 participating centers. Although CT was the predominant first imaging modality performed within one hour of emergency department arrival (83.6% overall), the proportion of MR-first workflows varied markedly across institutions, ranging from 1.0% to 56.7% (Figure 2). Notwithstanding the predominance of CT as the initial modality at most centers, a considerable proportion of patients underwent supplementary MR imaging within three hours of arrival, such that 64.6% (range, 30.0% to 81.0%) of all patients had received MR imaging within this interval.

**Figure 2.**
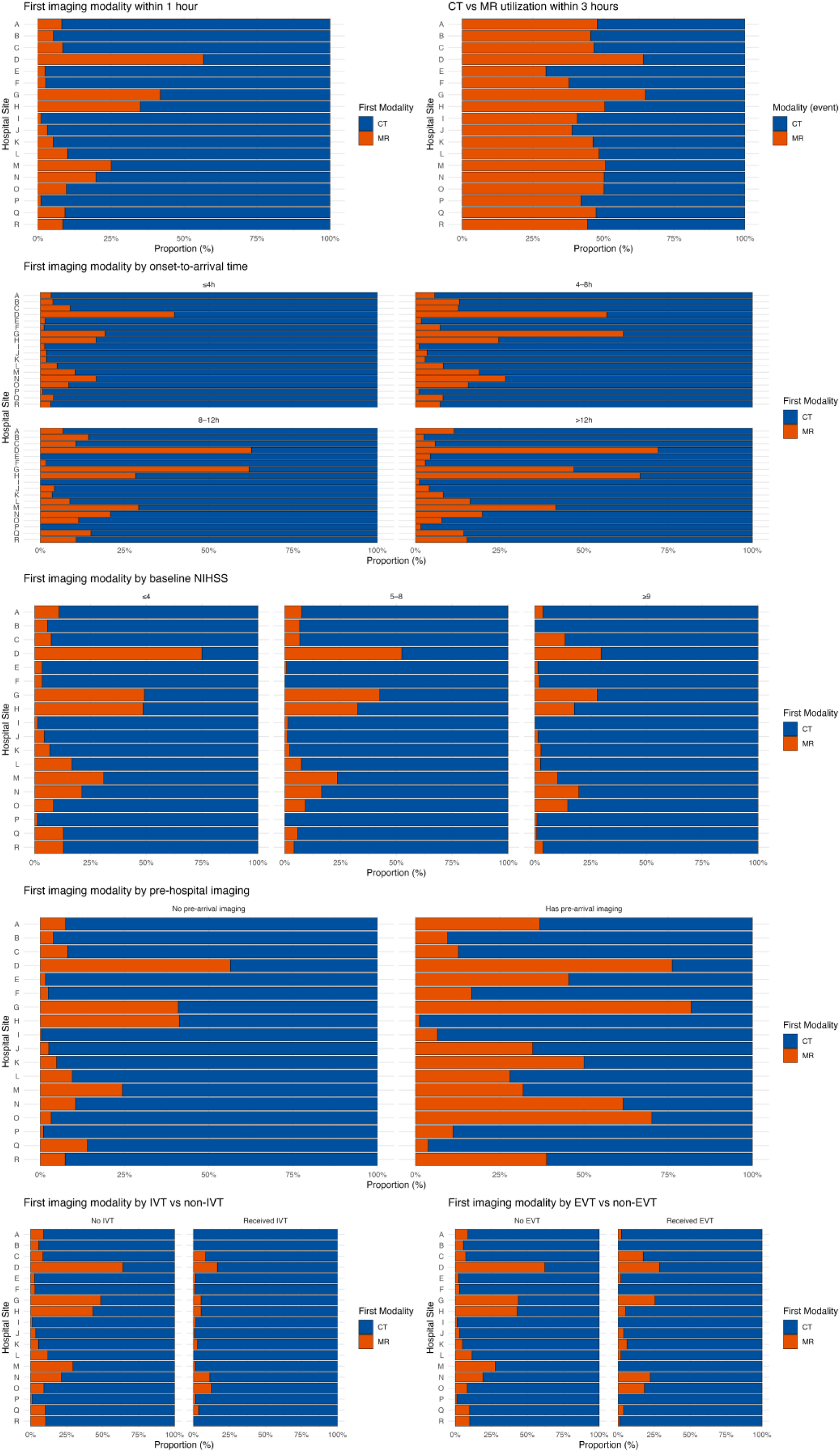
Inter-Hospital Variation in Acute Stroke Triaging Imaging Modality. Proportion of CT-based and MR-based imaging across 18 participating centers; Top row, first imaging modality in ER (left) and cumulative proportions of patients who had undergone CT or MR imaging within 3 hours after arrival; Second row, first imaging modality by last known well to arrival; Third row, first imaging modality by initial NIHSS score; Fourth row, first imaging modality by pre-hospital imaging; Bottom row, first imaging modality by intravenous or endovascular treatment. Abbreviations: CT, computed tomography; EVT, endovascular treatment; IVT, intravenous thrombolysis; MR, magnetic resonance; NIHSS, National Institutes of Health Stroke Scale.

MR-first workflows were increasingly prevalent with longer intervals from last known well to arrival (14.7% for 0 to 4.5 hours, 25.8% for 4.5 to 12 hours, 30.7% for 12 to 24 hours, and 44.6% for >24 hours; P for trend <0.01) and less frequent with increasing stroke severity (33.3% for NIHSS 0 to 5, 23.4% for 6 to 10, 16.6% for 11 to 15, 14.7% for 16 to 20, and 13.0% for >20; P for trend <0.01). Patients who arrived with pre-existing stroke imaging from a referring facility were substantially more likely to undergo MR as their first in-hospital study (52.5% versus 24.7%; P <0.01). Among patients who received recanalization treatment, CT-first workflows predominated, though EVT-treated patients exhibited a somewhat higher proportion of MR-first imaging than those receiving IVT (15.1% versus 7.7%; P <0.01); patients who received no recanalization treatment had the highest rate of MR-first workflows (32.4%).

### Acute Stroke Triaging Neuroimaging Workflow and Treatment Efficiency

Among patients who received IVT or EVT, those who had undergone additional imaging sequences prior to treatment tended to present with longer onset-to-arrival intervals and lower baseline NIHSS scores (Supplemental Tables S4 and S5). Across all the neuroimaging workflow groups, each additional imaging sequence category was associated with progressive prolongation of door-to-treatment time. Adjusted estimated mean door-to-needle times for IVT ranged from 28.6 ± 2.1 minutes for patients with pre-arrival imaging only to 79.3 ± 5.2 minutes for those who underwent MR with perfusion imaging prior to treatment; corresponding adjusted door-to-puncture times for EVT ranged from 52.4 ± 4.0 minutes to 139.2 ± 9.8 minutes (Table 2 and Figure 3).

**Figure 3.**
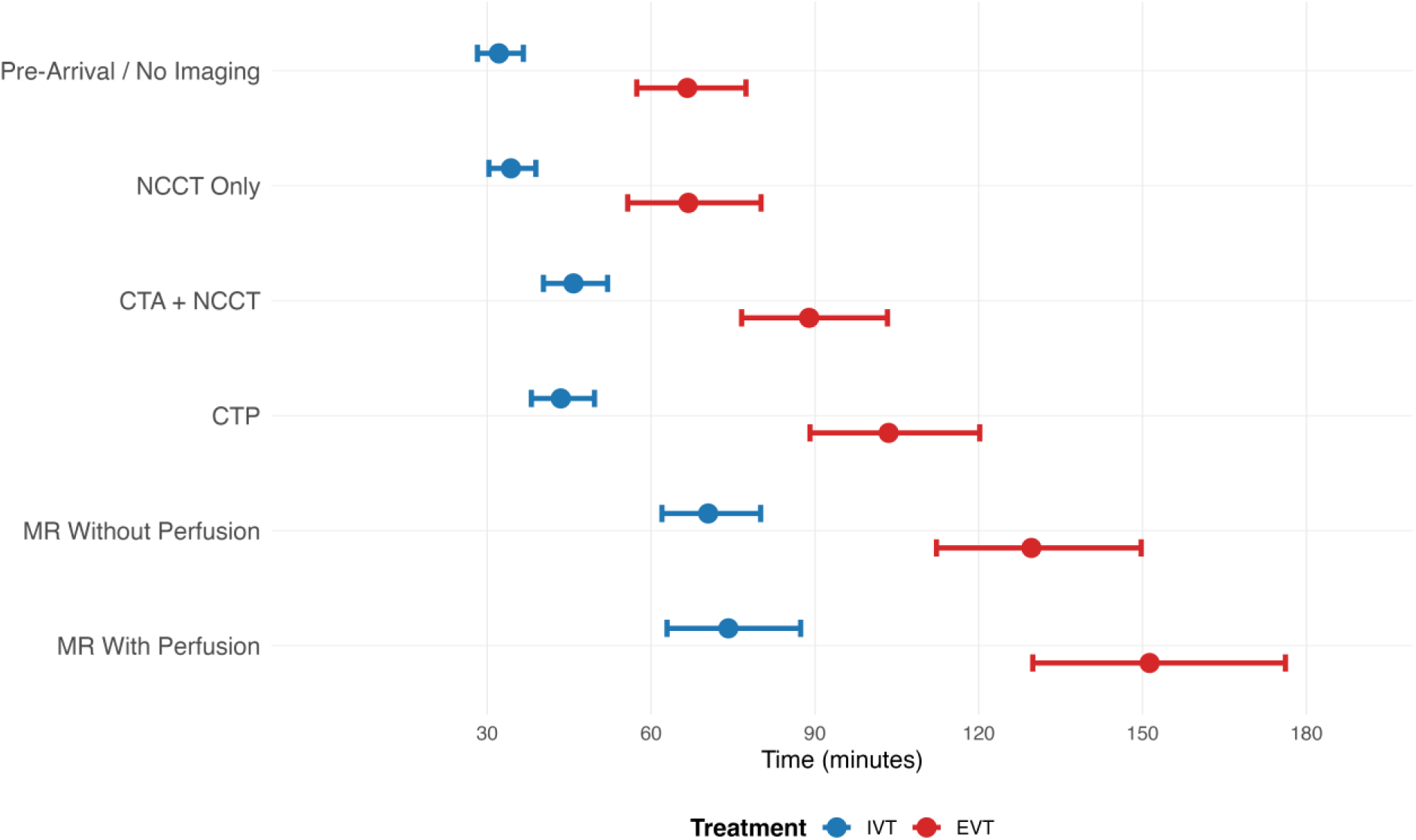
Adjusted Door-to-Treatment Time by Pre-Treatment Imaging Protocol. Estimated mean door-to-needle time (IVT; blue) and door-to-puncture time (EVT; red) across six hierarchically ordered imaging protocol categories. Estimates derived from multivariable linear mixed-effects models applied to log-transformed treatment times, adjusted for age, sex, onset-to-arrival time, baseline NIHSS, stroke etiology, vascular risk factors, and arrival year-month, with hospital as a random intercept; EVT models additionally adjusted for bridging thrombolysis. Error bars denote standard errors. Abbreviations: CTA, CT angiography; CTP, CT perfusion; EVT, endovascular treatment; IVT, intravenous thrombolysis; MR, magnetic resonance; NCCT, non-contrast computed tomography.

**Table 2.**
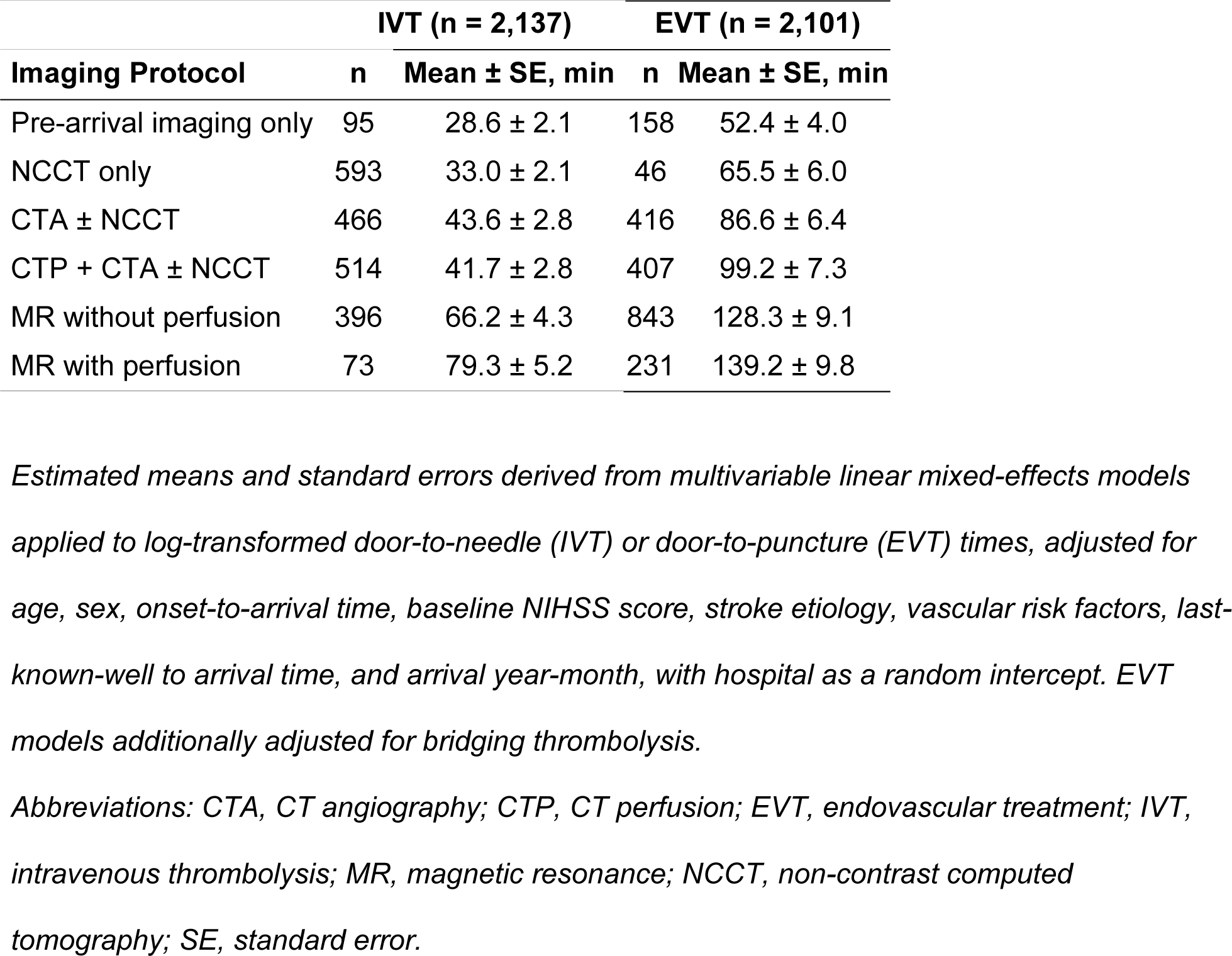
Adjusted Door-to-Treatment Times According to Pre-Treatment Imaging Protocol.

| Imaging Protocol | IVT (n = 2,137) |  | EVT (n = 2,101) |  |
| --- | --- | --- | --- | --- |
| | n | Mean ± SE, min | n | Mean ± SE, min |
| Pre-arrival imaging only | 95 | 28.6 ± 2.1 | 158 | 52.4 ± 4.0 |
| NCCT only | 593 | 33.0 ± 2.1 | 46 | 65.5 ± 6.0 |
| CTA ± NCCT | 466 | 43.6 ± 2.8 | 416 | 86.6 ± 6.4 |
| CTP + CTA ± NCCT | 514 | 41.7 ± 2.8 | 407 | 99.2 ± 7.3 |
| MR without perfusion | 396 | 66.2 ± 4.3 | 843 | 128.3 ± 9.1 |
| MR with perfusion | 73 | 79.3 ± 5.2 | 231 | 139.2 ± 9.8 |

Estimated means and standard errors derived from multivariable linear mixed-effects models applied to log-transformed door-to-needle (IVT) or door-to-puncture (EVT) times, adjusted for age, sex, onset-to-arrival time, baseline NIHSS score, stroke etiology, vascular risk factors, last-known-well to arrival time, and arrival year-month, with hospital as a random intercept. EVT models additionally adjusted for bridging thrombolysis.
Abbreviations: CTA, CT angiography; CTP, CT perfusion; EVT, endovascular treatment; IVT, intravenous thrombolysis; MR, magnetic resonance; NCCT, non-contrast computed tomography; SE, standard error.

### Proof-of-Concept Analysis: Initial Neuroimaging Modality and Benefit of Recanalization Treatment

Across both IVT and EVT cohorts, patients who underwent MR-based imaging before treatment initiation tended to have longer onset-to-arrival intervals and lower baseline NIHSS scores (Supplemental Tables S6 and S7).

Among EVT-treated patients, propensity score overlap-weighted analyses suggested numerically more favorable functional outcomes with CT-based imaging compared with MR-based imaging, particularly for dichotomized mRS outcomes (Table 3). In contrast, effect estimates among IVT-treated patients were closer to the null and exhibited greater variability across outcomes and analytic approaches. In sensitivity analyses, population-averaged marginal predicted risks estimated from propensity score-weighted logistic regression models were directionally consistent with the primary odds ratio-based findings (Supplemental Table S8). Among EVT-treated patients, MR-based imaging was associated with numerically lower predicted probabilities of favorable outcomes compared with CT-based imaging, whereas differences were smaller and less consistent among IVT-treated patients.

**Table 3.**
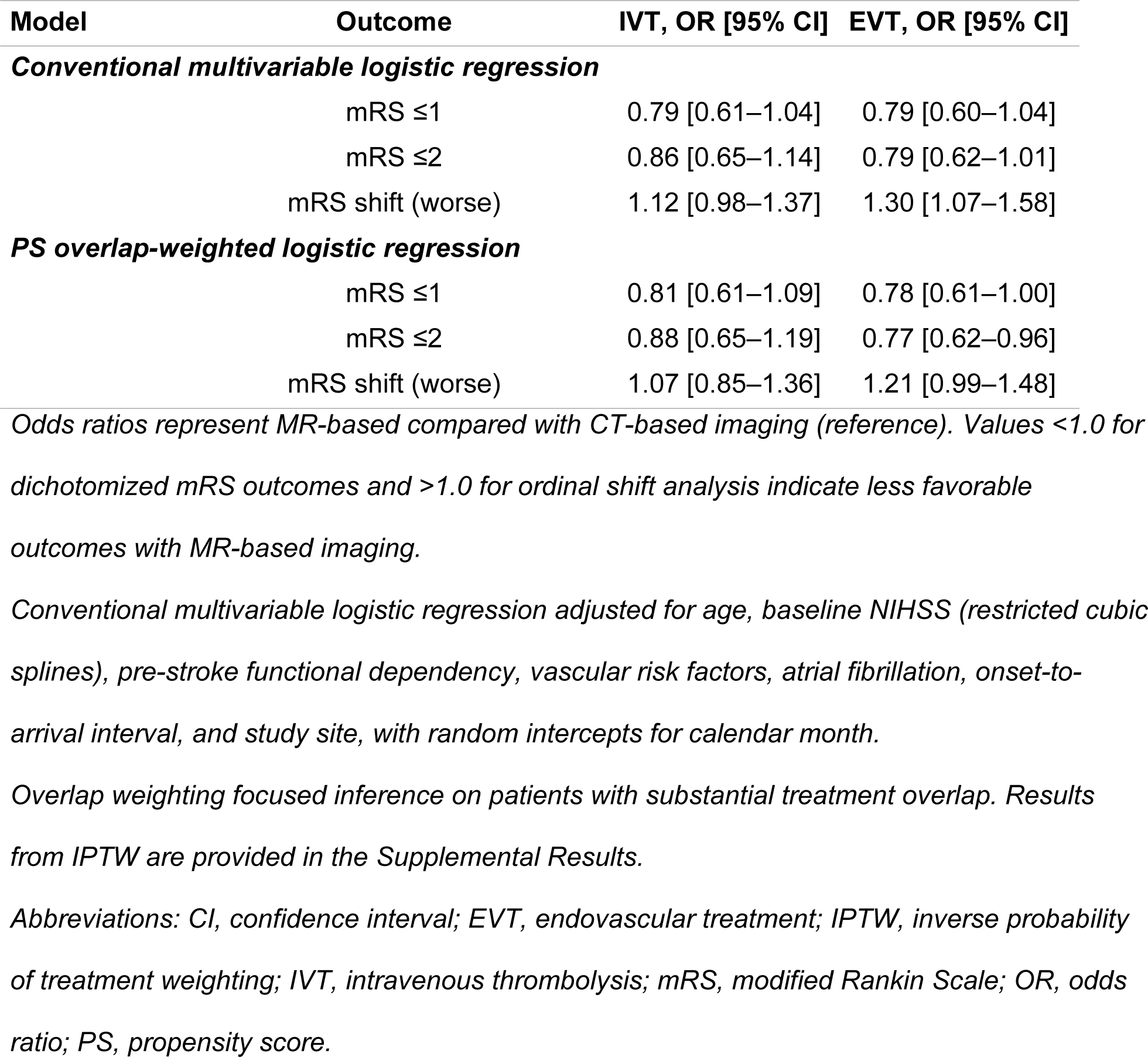
Associations Between MR-Based Compared With CT-Based Imaging Strategies and Functional Outcomes After Intravenous Thrombolysis or Endovascular Thrombectomy.

## Discussion

In this study, we delineate the development and current status of the CRCS-K Imaging Repository, a prospective multicenter neuroimaging repository that systematically collects all stroke neuroimaging studies during the index hospitalization and integrates them with clinical and outcome data. Over a three-year accrual period, the repository accumulated over 225,000 imaging sequences from 20,792 acute ischemic stroke patients across 18 comprehensive stroke centers. Leveraging AI-based automated quantification, the repository transforms routine clinical neuroimaging into standardized numeric features, disseminated through the AISCAN research platform. As a proof-of-concept application, we demonstrated substantial inter-hospital variation in imaging strategies, a systematic relationship between imaging modality and treatment timeliness, and numerically more favorable functional outcomes associated with CT-based protocols in EVT-treated patients.

Clinical registries have historically served as invaluable instruments for advancing stroke research, affording insights into real-world outcomes, healthcare disparities, and treatment effectiveness that randomized trials with restrictive eligibility criteria cannot readily provide.^25–27^ By collecting all neuroimaging in its original DICOM form, the CRCS-K Imaging Repository preserves the full dimensionality of imaging data for both contemporary and future analyses. Because imaging was collected at the individual sequence level rather than reduced to simplified categorical labels, we were able to characterize how acute stroke triage actually operates across 18 institutions, revealing substantial inter-hospital variation in modality selection that would be entirely invisible within conventional registry data. Importantly, this variation is not stochastic but rather systematically governed by clinical context: longer onset-to-arrival intervals and lower stroke severity shift clinicians toward MR-first workflows, whereas higher NIHSS scores and candidacy for acute recanalization treatment favor CT-first approaches. Of particular note, patients who arrived with imaging already obtained at a referring facility were substantially more likely to undergo MR as their initial in-hospital study, an observation with practical implications for the design of hub-and-spoke stroke networks, where the referring hospital’s imaging may fundamentally alter the receiving center’s workflow. These patterns reflect a pragmatic, clinician-driven decision process shaped by the interplay of time pressure, diagnostic uncertainty, and institutional resources, precisely the kind of real-world complexity that protocol-driven datasets cannot comprehensively capture.^28,29^

Beyond preserving imaging dimensionality, the repository addresses the qualitative, reader-dependent nature of conventional neuroimaging interpretation through AI-based automated quantification.^30^ Ischemic lesion volumes on NCCT and DWI, perfusion parameters, white matter hyperintensity burden, cerebral microbleed counts, and large vessel occlusion probability are converted into standardized numeric features applicable across the full imaging archive. Applied consistently across the full imaging archive, this approach reduces inter-rater variability and enables systematic, reproducible analysis at scale. The diagnostic and segmentation performance of several key modules has been independently validated in multicenter studies.^6,8,15–22^ Importantly, because the repository retains the original DICOM files, newly developed quantification algorithms in the future can be retrospectively applied to the entire archive without re-collection, an advantage that static, pre-quantified datasets cannot offer.

To disseminate the integrated dataset, we developed AISCAN, a web-based research platform that consolidates clinical, imaging, and AI-derived quantitative features within a unified research environment. Its architecture supports both centralized and standalone deployment, accommodating the heterogeneous privacy regulations across national and institutional jurisdictions. This design allows each participating site to maintain local governance of its data while operating within a shared analytical framework, establishing the potential for federated learning-based multinational research collaboration.^31–33^ The platform’s scalability has already been validated by its accommodation of the full 20,792-patient dataset and over 225,000 imaging sequences described herein.

Several imaging-enriched stroke registries have been described, each occupying a distinct niche. The MAGIC repository aggregates retrospective imaging from multiple European thrombectomy registries, offering a large sample of endovascular treatment cases with imaging selection data.^11^ The EVA-TRISP registry prospectively collects revascularization imaging across European centers.^12^ The CRCS-K Imaging Repository differs from these initiatives in three principal respects. First, it collects all neuroimaging performed during the entire index hospitalization, encompassing diagnostic, follow-up, and surveillance imaging beyond the acute treatment window, rather than restricting collection to treatment-relevant studies. Second, AI-based automated quantification is embedded within the data pipeline as a core architectural element rather than applied post hoc. Third, the registry is not restricted to patients who underwent recanalization treatment; the full spectrum of acute ischemic stroke severity and management is represented, enabling investigation of imaging practice patterns, treatment selection, and secondary prevention strategies alongside acute treatment efficacy. These features are counterbalanced by limitations: the geographic restriction to the Republic of Korea reduces ethnic and healthcare-system diversity, and the relatively high prevalence of MR-first workflows may limit direct comparability with CT-dominant systems. We utilized the integrated dataset to examine how pre-treatment imaging workflow composition affects treatment timeliness and functional outcome. Each additional imaging sequence was consistently associated with prolonged door-to-treatment time in both IVT and EVT, quantifying a trade-off that clinicians intuitively apprehend but that has rarely been measured at the sequence-level granularity. Propensity score analyses suggested numerically more favorable functional outcomes with CT-based imaging among EVT-treated patients; differences among IVT-treated patients were smaller and less consistent.

These findings, however, warrant cautious interpretation and should not be construed as evidence that CT-based imaging is inherently superior for treatment decision-making. The selection of imaging modality is powerfully confounded by indication: as demonstrated in our data, patients triaged with MR tended to present with lower NIHSS scores and longer onset-to-arrival intervals, a clinical profile reflective of diagnostic uncertainty rather than straightforward recanalization candidacy. Despite propensity score adjustment, residual confounding attributable to unmeasured factors, including clinical gestalt, fluctuating symptomatology, and institutional culture, cannot be excluded. Moreover, the time-outcome trade-off is intrinsic to this question: more comprehensive imaging incurs temporal cost but may confer more precise patient selection, and the net balance is likely contingent upon the clinical scenario. Landmark thrombectomy trials employed heterogeneous imaging protocols, and the optimal strategy for recanalization decision-making remains unresolved.^34–37^ The purpose of this demonstration was not to adjudicate the CT-versus-MR debate, but to illustrate how sequence-level imaging data, linked with outcomes, can generate pragmatic evidence that protocol-driven datasets are not designed to provide.^38^

Several limitations merit consideration. With respect to the repository itself, the collected imaging modalities are confined to CT, MR, and angiography; transcranial Doppler and carotid ultrasonography were not included owing to non-standardized institutional protocols. The participating centers comprise academic comprehensive stroke centers in the Republic of Korea, and generalizability to community hospitals or other healthcare systems warrants circumspection. Consent-dependent enrollment may engender underrepresentation of patients with severe stroke who are unable to provide consent and lack available legal representatives, although the 92% capture rate among eligible patients during the study period mitigates this concern to a considerable degree.^13^

Regarding the demonstration analysis, limitations beyond the confounding by indication discussed above include the following. The three-hour threshold adopted to define acute triaging imaging is inherently arbitrary, and alternative temporal cutoffs may yield different classifications. Finally, MR-first workflows are substantially more prevalent in the Republic of Korea and selected East Asian countries than in most Western healthcare systems, which may circumscribe the international generalizability of the workflow analysis findings.^11,12^ Conversely, this very characteristic constitutes a distinctive research advantage, as the coexistence of CT-dominant and MR-dominant institutions within a single registry provides a natural laboratory for the influence of imaging modalities on the efficiency and outcomes.

The CRCS-K Imaging Repository and its research platform AISCAN bridge the divide between clinical stroke registries and the multidimensional information embedded in routine neuroimaging. Its prospective, continuously maintained design accommodates the evolution of clinical practice and emerging AI tools, providing durable infrastructure for longitudinal investigation. The initial application presented herein illustrates the breadth of questions that become addressable when granular imaging and clinical data coexist within a queryable framework. Future applications include cross-national benchmarking of imaging practice, observational emulation of trial designs, and longitudinal assessment of how AI deployment transforms clinical workflows. We envision this repository as a model for how prospective stroke registries can evolve into dynamic, image-rich research ecosystems that advance in concert with the integration of artificial intelligence into cerebrovascular care.

